# Impact of stopping therapy during the SARS-CoV-2 pandemic in persons with lymphoma

**DOI:** 10.1101/2020.09.28.20203083

**Authors:** Shenmiao Yang, Dong Dong, Hongfei Gu, Robert Peter Gale, Jun Ma, Xiaojun Huang

## Abstract

**Introduction:** The severe acute respiratory syndrome-2 (SARS-CoV-2) pandemic disrupted medical care for persons with cancer including those with lymphoma. Many professional societies recommend postponing, decreasing or stopping anti-cancer therapy in selected persons during the pandemic. However, although seemingly sensible these recommendations are not evidence-based and their impact on anxiety and health-related *quality-of-life* (HRQoL) is unknown.

**Methods:** Surveyed 2532 subjects including 1060 persons with lymphoma, 948 caregivers and 524 normal, uninvolved persons using a purposed-designed questionnaire on a patient organization website. Respondents also completed the Zung Self-Rating Anxiety and patient respondents, the EORTC QLQ-C30 instruments to quantify anxiety and HRQoL. We also evaluated caregiver support and an online education programme of the Chinese Society of Clinical Oncology (CSCO). Data of HRQoL from a 2019 pre-pandemic online survey of 1106 persons with lymphoma was a control.

**Results:** 33% (95% Confidence Interval [CI] 30, 36%) of lymphoma patients and 31% (28, 34%) of caregivers but only 21% (17, 24%) of normal individuals had any level of anxiety (both pair-wise *P* < 0.001). Amongst lymphoma respondents physical exercise and better caregiver support were associated with less anxiety whereas female sex, receiving therapy and reduced therapy intensity were associated with more anxiety. Paradoxically, lymphoma respondents during the pandemic had better HRQoL than pre-pandemic controls. Reduced therapy intensity was associated with worse HRQoL whereas respondents who scored caregiver support and the online patient education programme high had better HRQoL.

**Conclusions:** During the SARS-CoV-2 pandemic lymphoma patients and their caregivers had significantly higher incidence of anxiety compared with normals. Lymphoma respondents reported better HRQoL compared with pre-pandemic controls. Reduced therapy-intensity in patients with cancer may have unanticipated adverse effects on anxiety and HRQoL. Regular and intense support by caregivers and online education programmes alleviate anxiety and improve HRQoL.

## Introduction

The severe acute respiratory virus coronavirus-2 (SARS-CoV-2) pandemic caused major changes also in health care support for patients with cancer worldwide (Maringe, et al. 2020; Shah, et al.; Sud, et al. 2020; van de Haar, et al. 2020). Recommendations from many professional societies suggest an individual based decision (Al-Shamsi, et al. 2020; Di Ciaccio, et al. 2020; Ismael, et al. 2020). The general thrust is to reduce therapy intensity in patients with cancer with concerns that cancer and treatment may lead to a higher risk of infections and worse coronavirus infectious disease-2019 (COVID-19) outcomes.

We selected lymphoma as a frequent type of cancer and sought to investigate how the pandemic and typical interventions might impact on levels of anxiety among patients and their caregivers as well as how they affected patients’ health related quality of life (HRQoL). Therefore, we did an online survey using the platform of the Chinese lymphoma patient organization (House086). Patients and caregivers completed a questionnaire and standardized evaluation instruments to quantify levels of anxiety and HRQoL. The questionnaire and anxiety instrument were also completed by normal individuals using the WeChat, a messaging and social media mobile app platform frequently used in China. We found the prevalence of anxiety in lymphoma patient and caregiver respondents during the SARS-CoV-2 pandemic was significantly higher compared with normal individuals. Unexpectedly, patient HRQoL during the pandemic was better compared with a propensity score matched pre-pandemic cohort. Better caregiver support was associated with less anxiety and better HRQoL. Access to an internet-based lymphoma patient support platform and an education programme improved HRQoL. Several of these co-variates are actionable and may help to alleviate patients’ concerns caused by SARS-CoV-2.

## Materials and methods

### Study Participants and Study Conduct

We conducted a cross-sectional survey of lymphoma patients and their caregivers regarding the level of anxiety and patients’ HRQoL between 17-19^th^ April, 2020, using House086 as the distribution platform (Additional file 1). Controls for the anxiety instrument were persons with no association with lymphoma patients or hospitals invited to participate in an online WeChat survey. Data from a HRQoL nationwide cross-sectional survey of lymphoma patients in 2019 were used to find matching cases as a pre-pandemic control cohort. The 2019 lymphoma survey included 4068 Chinese with all sub-types of lymphoma from which 1106 patients matched on sex, age, education level, and sub-type of lymphoma and therapy type were extracted.

Sample size was calculated as *n*= (*z*)^2^ *p* (1 – *p*) / *d*^2^, in which *z* = 1.96 for a level (α) of confidence of 95%. Tolerated margin of error was 0.05. The prevalence of clinically important anxiety in the Chinese population is reported as 29 to 35 percent (Huang and Zhao 2020; Wang, et al. 2020). Minimum number of the qualified questionnaires was estimated as 350.

An online questionnaire collected data on: (1) demographics; (2) lymphoma-related data; (3) impact of the pandemic on health care related activities; (4) hours of mobile phone use; (5) online patient-assistance resources; (6) quality of caregiver support (10-point scale); and (7) quality of the CSCO online education programme (10-point scale). To quantify anxiety of lymphoma patients, caregivers and normal individuals we used the Zung Self-Rating Anxiety Scale (SAS) (Chinese version) (Liu, et al. 1997; Zung 1971; ZY 1984). We used the EORTC QLQ-C30 (v.3; Chinese version) to quantify lymphoma patients’ HRQoL (Aaronson, et al. 1993; Zhao and Kanda 2000).

The study was approved by the Ethics Committees of Peking University Peoples’ Hospital according to tenets of the Declaration of Helsinki (Register number 2020PHB173). Electronic informed consent was obtained from all respondents who could withdraw at any time during the survey without prejudice.

### Statistical Analysis

Descriptive statistics were used for the demographic, social and lymphoma related co-variates. Anxiety index (AI) was calculated according to the Zung SAS, a rating instrument for anxiety disorders. Based on extensive validated data an AI < 50 is defined as *normal*, 50 to 59, minimal/moderate, 60 to 69, marked/severe and ≥ 70, extreme anxiety in the Chinese population with an internal consistency reliability of 0.66–0.80 and the Cronbach α of 0.87 (Minglu, et al. 2020; Shao, et al. 2020). Scores of five functioning scales (physical, role, emotional, cognitive, and social functioning), eight symptom scales (fatigue, nausea/vomiting, pain, dyspnea, sleep disturbances, appetite loss, constipation, and diarrhea), fiscal impact and overall HRQoL were calculated using the EORTC QLQ-C30 instrument (Aaronson, et al. 1993).

Lymphoma respondents from this study were matched with respondents to the 2019 pre-pandemic study on co-variates including age, education level, lymphoma sub-type, and therapy by the nearest neighbor matching method with R packages “MatchIt” at a 1:1 ratio. Standardized mean difference was calculated for each of the co-variates between the cohorts before and after matching to assess matching quality. An absolute standardized difference of > 20% denotes meaningful co-variate imbalance. (Additional file 2)

We used the Independent-Samples t-test to compare groups of continuous variables and 1-way analysis of variance (ANOVA) and LSD to analyze differences among cohorts and each paired cohort. Chi-square test was used to analyze categorical co-variates. We used a multi-variable analysis with binary logistic regression to identify risk factors of anxiety. The Kendall tau-b correlation was used to evaluate risk factors of HRQoL. Tests were 2-sided and *P* ≤ 0.05 was considered significant. Statistical analyses were performed with SPSS 12.0 (IBM SPSS Statistics, New York, US).

## Results

We received 2745 responses from subjects in 32 Chinese provinces, autonomous regions, centrally-administered municipalities and special administrative regions identified by internet protocol (IP) addresses. 166 questionnaires, incomplete or completed in < 1 min or > 60 min, were excluded from further analyses. 94 percent of questionnaires were evaluable.

### Respondent co-variates

1106 (43%) of the 2578 respondents were lymphoma patients, 948 (37%), caregivers and 524 (20%), normal individuals (Table 1). 1031 respondents (40%) were male and 2313 (90%), 20 to 60 years. 495 (45%) patient respondents were 20 to 39 years, 477 (43%) were 40 to 59 years and 110 (10%), ≥ 60 years. There was a discordance between patients’ age reported by patients and caregivers (Table 2). 1912 (74%) respondents were college graduates or received other higher education. 88 (3%) lived in Hubei province with Wuhan as the capital city known to be the first area on lockdown from 23^rd^ January to 8^th^ April in China. 275 (11%) were living outside their usual residence. 15 reported they were infected with SARS-CoV-2 and another 19, their friends or relatives were infected with SARS-CoV-2.

**Table 1.** Co-variates of all respondents.

|  | No. (%) |  |  |
| --- | --- | --- | --- |
|  | Patients (*n*=1106) | Caregivers (*n*=948) | Normal individuals (*n*=524) |
| Male | 482 (44) | 293 (31) | 256 (49) |
| Age, y |  |  |  |
| 18-20 | 24 (2) | 8 (1) | 70 (13) |
| 20-39 | 495 (45) | 544 (57) | 280 (53) |
| 40-59 | 477 (43) | 371 (39) | 146 (28) |
| 60-79 | 108 (10) | 25 (3) | 27 (5) |
| ≥ 80 | 2 (1) | 0 | 1 (1) |
| Education |  |  |  |
| Primary school | 25 (2) | 18 (2) | 6 (1) |
| Middle/High school | 331 (30) | 195 (21) | 91 (17) |
| College/University | 682 (62) | 639 (67) | 379 (72) |
| ≥ Postgraduate | 68 (6) | 96 (10) | 48 (9) |
| SARS-COV-2-infection | 5 (1) | 10 (1) | 0 |
| SARS-CoV-2-infection in family or friends | 5 (1) | 11(1) | 3 (1) |
| Hubei resident | 52 (5) | 30 (3) | 6 (1) |
| Physical exercise |  |  |  |
| No | 214 (19) | 251 (27) | 130 (25) |
| Yes | 892 (80) | 697 (74) | 394 (75) |
| Mobile phone use pre-pandemic, hours/day |  |  |  |
| < 3 | 551 (50) | 549 (58) | 268 (51) |
| 4-5 | 361 (33) | 263 (28) | 157 (31) |
| 6-7 | 126 (11) | 90 (10) | 57 (11) |
| ≥ 8 | 68 (6) | 46 (5) | 42 (8) |
| Mobile phone hours/day during pandemic |  |  |  |
| < 3 | 358 (32) | 308 (33) | 149 (28) |
| 4-5 | 409 (37) | 376 (40) | 181 (35) |
| 6-7 | 204 (18) | 167 (18) | 98 (19) |
| ≥ 8 | 135 (12) | 97 (10) | 96 (18) |
| Concerns |  |  |  |
| SARS-CoV-2-infection | 547 (50) | 392 (36) | 308 (59) |
| SARS-CoV-2 infection of family/ friends | 214 (19) | 146 (15) | 347 (66) |
| Family separation | 28 (3) | 22 (2) | 180 (34) |
| Income loss | 204 (18) | 151 (16) | 198 (38) |
| Lymphoma | 603 (55) | 595 (63) | - |
| Read online or obtained information |  |  |  |
| SARS-CoV-2 infection or/and COVID-19 | 946 (86) | 822 (87) | 471 (90) |
| Lymphoma | 572 (84) | 539 (91) | - |
| Entertainment | 610 (55) | 370 (39) | 410 (78) |
| Work | 209 (19) | 228 (24) | 252 (48) |
| Family support |  |  |  |
| Score, mean (SD) | 8.84 (2.16) | 8.93 (1.97) | 8.25 (2.10) |
| Anxiety index, mean (SD) | 45.8 (9.4) | 46.0 (9.8) | 42.6 (9.9) |
| Clinical interpretation of AI |  |  |  |
| Normal (< 50) | 793 (67) | 654 (69) | 416 (79) |
| Minimal/moderate (50-59) | 284 (26) | 207 (22) | 78 (15) |
| Marked/ever anxiety (60-69) | 70 (6) | 67 (7) | 24(5) |
| Extreme ($\geq 70$) | 13 (1) | 20 (2) | 6 (1) |

**Table 2.** Co-variates of lymphoma patient respondents.

|  | No. (%) |  |  |
| --- | --- | --- | --- |
|  | Reported by patients (*n*=1106) | Reported by caregivers (*n*=948) | Both (*n*=2054) |
| Male | 482 (44) | 541 (57) | 1023 (50) |
| Age, y |  |  |  |
| <20 | 24 (2) | 96 (10) | 120 (6) |
| 20-39 | 495 (45) | 184 (19) | 679 (33) |
| 40-59 | 477 (43) | 368 (39) | 845 (41) |
| 60-79 | 108 (10) | 291 (31) | 399 (19) |
| ≥80 | 2 (1) | 9 (1) | 11 (1) |
| Lymphoma type |  |  |  |
| Hodgkin | 89 (8) | 70 (7) | 159 (8) |
| Non-Hodgkin |  |  |  |
| Indolent | 361 (33) | 174 (18) | 535 (26) |
| Follicular | 210 (19) | 86 (9) | 296 (14) |
| Marginal zone | 64 (6) | 39 (4) | 103 (5) |
| CLL/SLL | 69 (6) | 31 (3) | 100 (5) |
| Lymphoplasmacytic | 3 (1) | 3 (1) | 6(1) |
| Other | 15 (1) | 15 (2) | 30 (2) |
| Aggressive | 654 (59) | 700 (74) | 1354 (66) |
| Diffuse large B-cell | 405 (37) | 413 (44) | 818 (40) |
| Burkitt | 26 (2) | 30 (3) | 56 (3) |
| Mantle cell | 30 (3) | 39 (4) | 69 (3) |
| Extra-nodal NK/T-cell | 66 (6) | 61 (6) | 127 (6) |
| Peripheral T-cell | 32 (3) | 46 (5) | 78 (4) |
| Anaplastic large T-cell | 43 (4) | 46 (5) | 89 (4) |
| Lymphoblastic | 37 (3) | 46 (5) | 83 (4) |
| Other T-cell | 15 (1) | 19 (2) | 34 (2) |
| Unknown | 2 (1) | 4 (1) | 6 (1) |
| Stage |  |  |  |
| Early | 416 (38) | 279 (29) | 695 (34) |
| Advanced | 519 (53) | 556 (59) | 1075 (52) |
| Unknown | 99 (9) | 113 (12) | 212 (10) |
| On-therapy |  |  |  |
| No | 676 (61) | 412 (44) | 1088 (53) |
| Parenteral | 301 (27) | 406 (43) | 707 (34) |
| Oral | 129 (12) | 130 (14) | 259 (13) |
| Changed hospital | 633 (57) | 522 (55) | 1155 (56) |
| Change in medical activities |  |  |  |
| Delayed therapy | 124 (11) | 135 (14) | 259 (13) |
| Less intensive therapy | 82 (7) | 110 (12) | 192 (9) |
| Switched to oral drugs | 41 (4) | 48 (5) | 89 (4) |
| Delayed exams | 441 (40) | 317 (33) | 728 (35) |
| No change | 570 (52) | 489 (52) | 1059 (52) |
| Reduced-intensity | 217 (20) | 265 (28) | 482 (24) |
| Result |  |  |  |
| Assistance in confirming lymphoma diagnosis | 208 (31) | 215 (36) | 423 (33) |
| Therapy assistance | 246 (36) | 271 (46) | 517 (41) |
| SARS-CoV-2 prevention or COVID-19 therapy | 163 (24) | 133 (23) | 296 (23) |
| Physician communication | 201 (29) | 195 (33) | 396 (31) |
| Patient/caregiver communication | 226 (33) | 222 (38) | 448 (35) |
| Improved mood | 325 (48) | 278 (47) | 603 (47) |
| Increased confidence | 362 (53) | 372 (53) | 734 (58) |

Distribution of lymphoma types reported by patient and caregiver respondents was similar to lymphoma distribution data from China (Cao, et al. 2018; Sun, et al. 2012; Yang, et al. 2011). 654 (59%) patient and 700 (74%) caregiver respondents reported an aggressive lymphoma (*P* <0.001; Table 2). 966 (47%) patient respondents were on-therapy, parenteral in 707 (34%) and oral in 259 (13%). 1088 (53%) patient respondents were under medical supervision with no current therapy. 819 (47%) receiving in-hospital (40%) or outpatient (7%) therapy.

1155 (56%) patients changed the hospital they had been visiting for routine monitoring and/or therapy during the SARS-CoV-2 pandemic. 192 (9%) changed to a therapy of lower intensity. 89 (4%) switched to oral anti-lymphoma drugs, 259 (13%) delayed scheduled parenteral therapy and 761 (37%) delayed or postponed scheduled hospital visits. 482 (24%) experienced reduced therapy intensity including fewer drugs, reduced drug doses, a switch from parenteral to oral drugs and/or therapy delay or discontinuation. 1059 (52%) reported no change of their medical activities including physician visits, exams and/or therapy.

### Respondent Concerns

The most frequent concerns of patient respondents were their lymphoma (*N* = 603; 55%), SARS-CoV-2-infection (*N* = 547; 50%) and the inability to attend outpatient clinics (N = 429; 39%). The most frequent concerns for caregiver respondents related to the patient they were caring for were lymphoma (*N* = 595; 63%), therapy-disruption (*N* = 397; 42%) and SARS-CoV-2-infection (*N* = 392; 41%). The most frequent concerns of normal respondents were SARS-CoV-2-infection risk to their family (*N* = 347; 66%), themselves (*N* = 308; 59%) and income loss (*N* = 198; 38%).

### Respondent Anxiety

Respondents with any level of anxiety (*i*.*e*. Zung score > 50) were 33% (95% Confidence Interval [CI] 30, 36%) for lymphoma patients, 31% (28, 34%) for caregivers and 21% (17, 24%; three-cohort comparison: *P* < 0.001) for normal individuals. Pair-wise comparisons showed incidence of anxiety was similar in patients and caregiver respondents (*P* = 0.29) but higher compared with normal individuals (both *P* < 0.001). Severity of anxiety was similar in the three cohorts. Among persons with anxiety, minimal/moderate severity levels (score > 49) were 77%, 70% and 72%, marked/severe anxiety (score > 59, 19%, 23% and 22% and extreme anxiety (score > 69), 4%, 7% and 6% (*P* = 0.22).

We evaluated co-variates associated with anxiety in respondents (Table 3). No SARS-CoV-2 infection (Hazard Ratio [HR] = 0.15 (0.041, 0.53; *P* = 0.003), being a normal *versus* a patient or caregiver (HR =0.53 [0.42, 0.68]; *P* < 0.001), physical exercise (HR = 0.60 (0.49, 0.73; *P* < 0.001), higher education level (HR = 0.77 (0.63, 0.93; *P* = 0.007), increase of > 2 hours/day of mobile phone use (HR = 0.83 [0.70, 0.99]; *P* = 0.042) and higher family support score (HR=0.93 [0.89, 0.96]; *P* = 0.001) were associated with less anxiety.

**Table 3.**
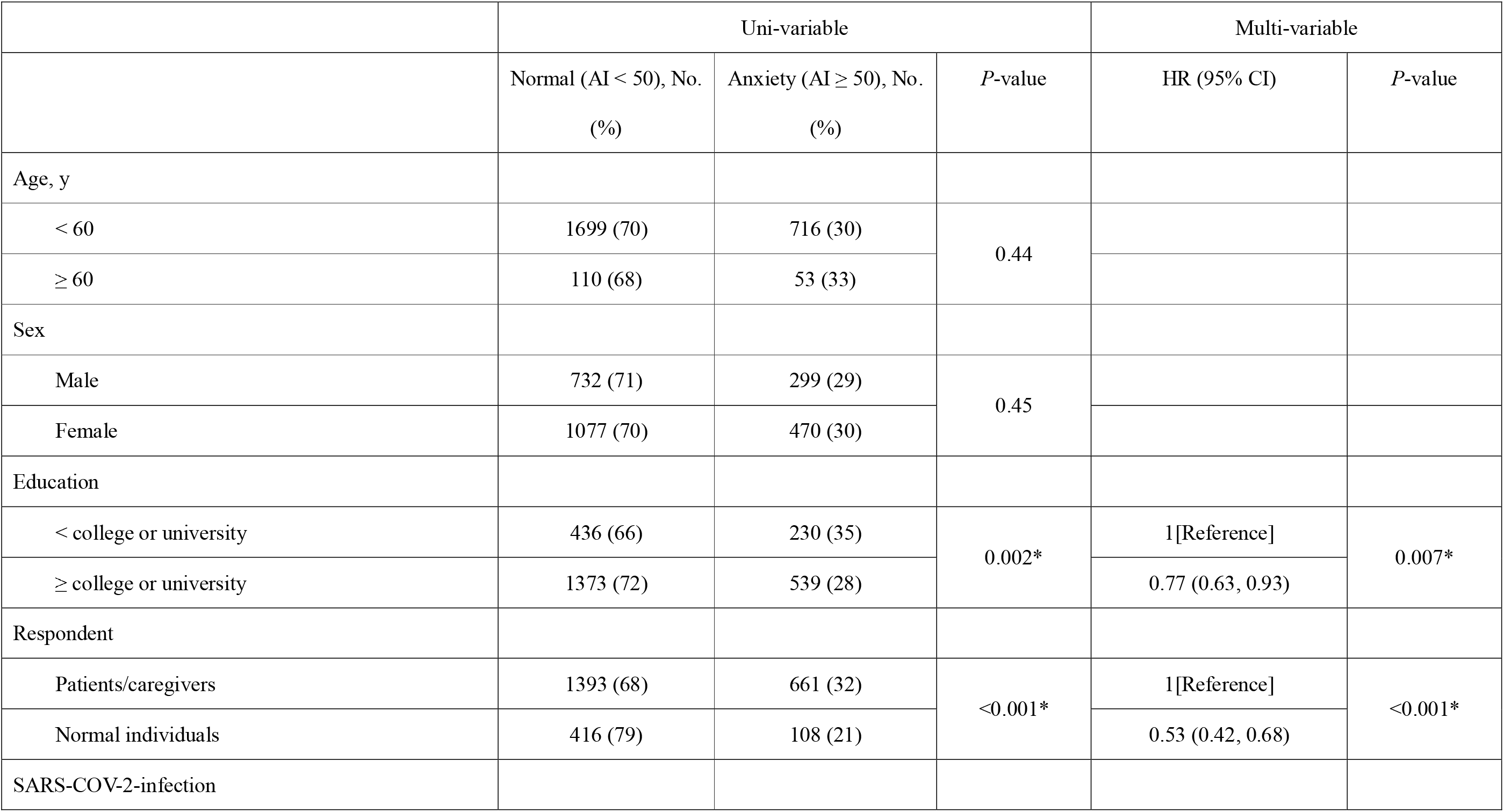
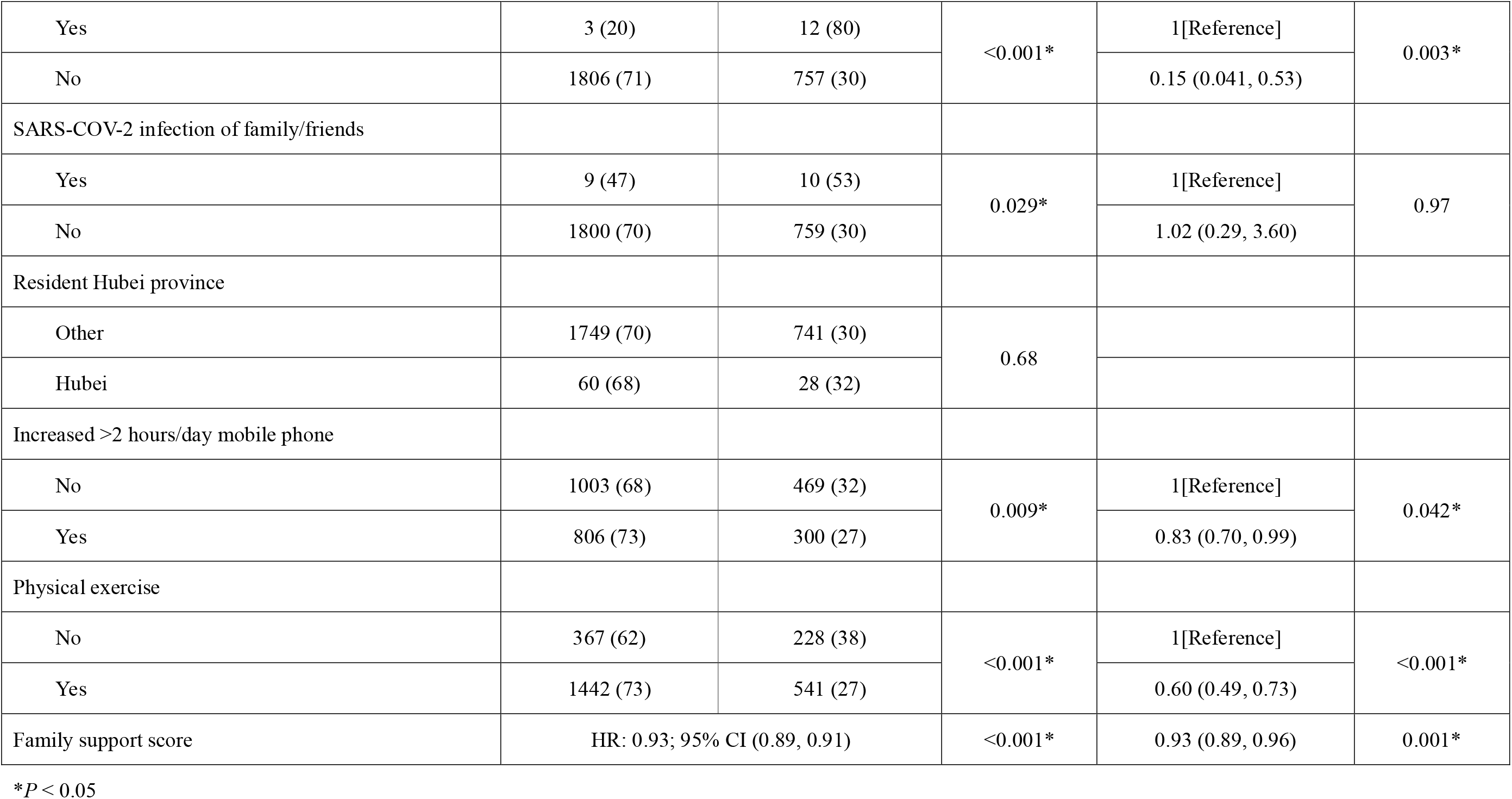
Risk factors of anxiety in all respondents.

|  | Uni-variable |  |  | Multi-variable |  |
| --- | --- | --- | --- | --- | --- |
|  | Normal (AI < 50), No.<br>(%) | Anxiety (AI ≥ 50), No.<br>(%) | *P*-value | HR (95% CI) | *P*-value |
| Age, y |  |  |  |  |  |
| < 60 | 1699 (70) | 716 (30) | 0.44 |  |  |
| ≥ 60 | 110 (68) | 53 (33) |  |  |  |
| Sex |  |  |  |  |  |
| Male | 732 (71) | 299 (29) | 0.45 |  |  |
| Female | 1077 (70) | 470 (30) |  |  |  |
| Education |  |  |  |  |  |
| < college or university | 436 (66) | 230 (35) | 0.002* | 1[Reference] | 0.007* |
| ≥ college or university | 1373 (72) | 539 (28) |  | 0.77 (0.63, 0.93) |  |
| Respondent |  |  |  |  |  |
| Patients/caregivers | 1393 (68) | 661 (32) | <0.001* | 1[Reference] | <0.001* |
| Normal individuals | 416 (79) | 108 (21) |  | 0.53 (0.42, 0.68) |  |
| SARS-COV-2-infection |  |  |  |  |  |
| Yes | 3 (20) | 12 (80) | <0.001* | 1[Reference] | 0.003* |
| No | 1806 (71) | 757 (30) |  | 0.15 (0.041, 0.53) |  |
| SARS-COV-2 infection of family/friends |  |  |  |  |  |
| Yes | 9 (47) | 10 (53) | 0.029* | 1[Reference] | 0.97 |
| No | 1800 (70) | 759 (30) |  | 1.02 (0.29, 3.60) |  |
| Resident Hubei province |  |  |  |  |  |
| Other | 1749 (70) | 741 (30) | 0.68 |  |  |
| Hubei | 60 (68) | 28 (32) |  |  |  |
| Increased >2 hours/day mobile phone |  |  |  |  |  |
| No | 1003 (68) | 469 (32) | 0.009* | 1[Reference] | 0.042* |
| Yes | 806 (73) | 300 (27) |  | 0.83 (0.70, 0.99) |  |
| Physical exercise |  |  |  |  |  |
| No | 367 (62) | 228 (38) | <0.001* | 1[Reference] | <0.001* |
| Yes | 1442 (73) | 541 (27) |  | 0.60 (0.49, 0.73) |  |
| Family support score | HR: 0.93; 95% CI (0.89, 0.91) |  | <0.001* | 0.93 (0.89, 0.96) | 0.001* |

\* $P < 0.05$

Patient respondents not hospitalized during the pandemic but before the survey had less anxiety of any severity compared with hospitalized respondents (HR = 0.62 [0.48, 0.81]; *P* = 0.004). Frequency of marked/severe or extreme severity was also increased in hospitalized *versus* not hospitalized patient respondents (HR = 0.62 [0.40, 0.98]; *P* = 0.042). Amongst patient respondents not hospitalized during the pandemic but before the survey, persons with a higher family support score had lower incidence of anxiety (HR = 0.90 [0.85, 0.96]; *P* = 0.002). Amongst patient respondents hospitalized during the pandemic but before the survey there was no correlation between incidence of anxiety and caregiver support score (HR= 0.97 [0.86, 1.08]; *P* = 0.54).

In multi-variable analyses, we found more patients receiving therapy (HR = 1.43 [1.08, 1.89]; *P* = 0.012), those with reduced intensity of therapy (HR = 1.59 [1.14, 2.21]; *P* = 0.006) and females (HR = 1.33 [1.02, 1.72]; *P* = 0.034) had a higher incidence of anxiety whereas physical exercise (HR = 0.57 [0.42, 0.77]; *P* < 0.001) and higher family support score (HR = 0.92 [0.87, 0.98]; *P* = 0.006) were associated with lower incidences of anxiety (Table 4).

**Table 4.**
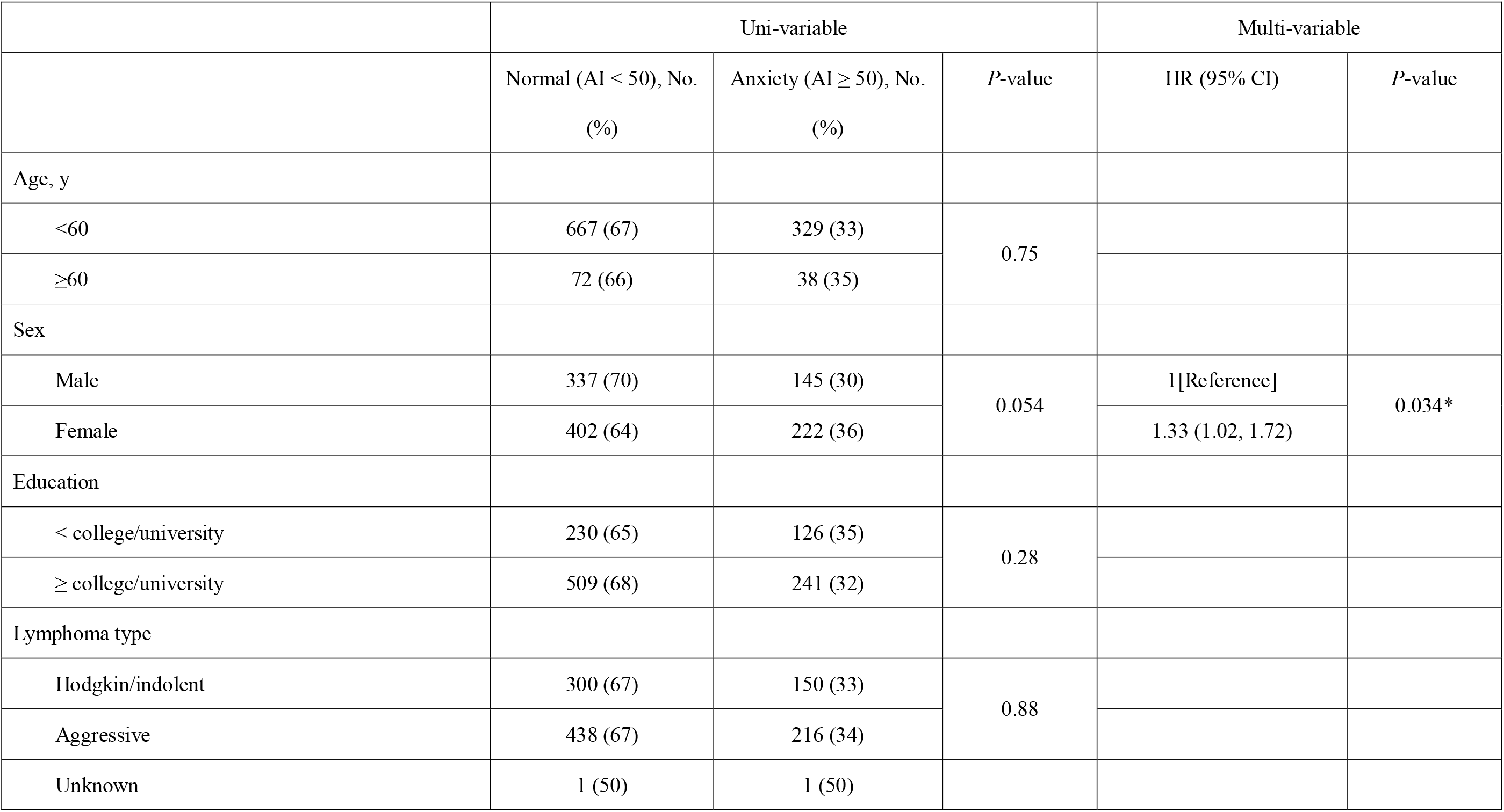
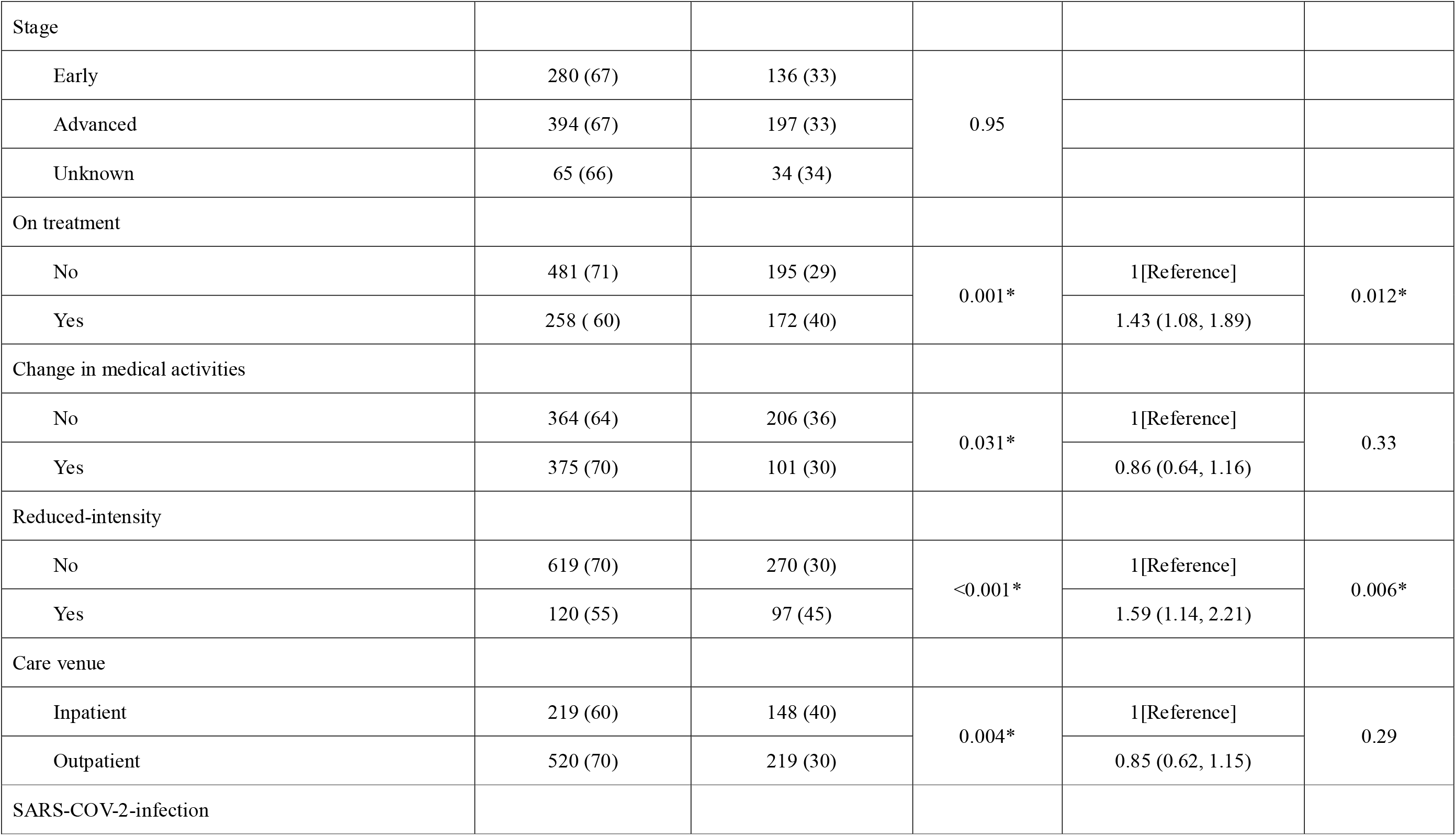
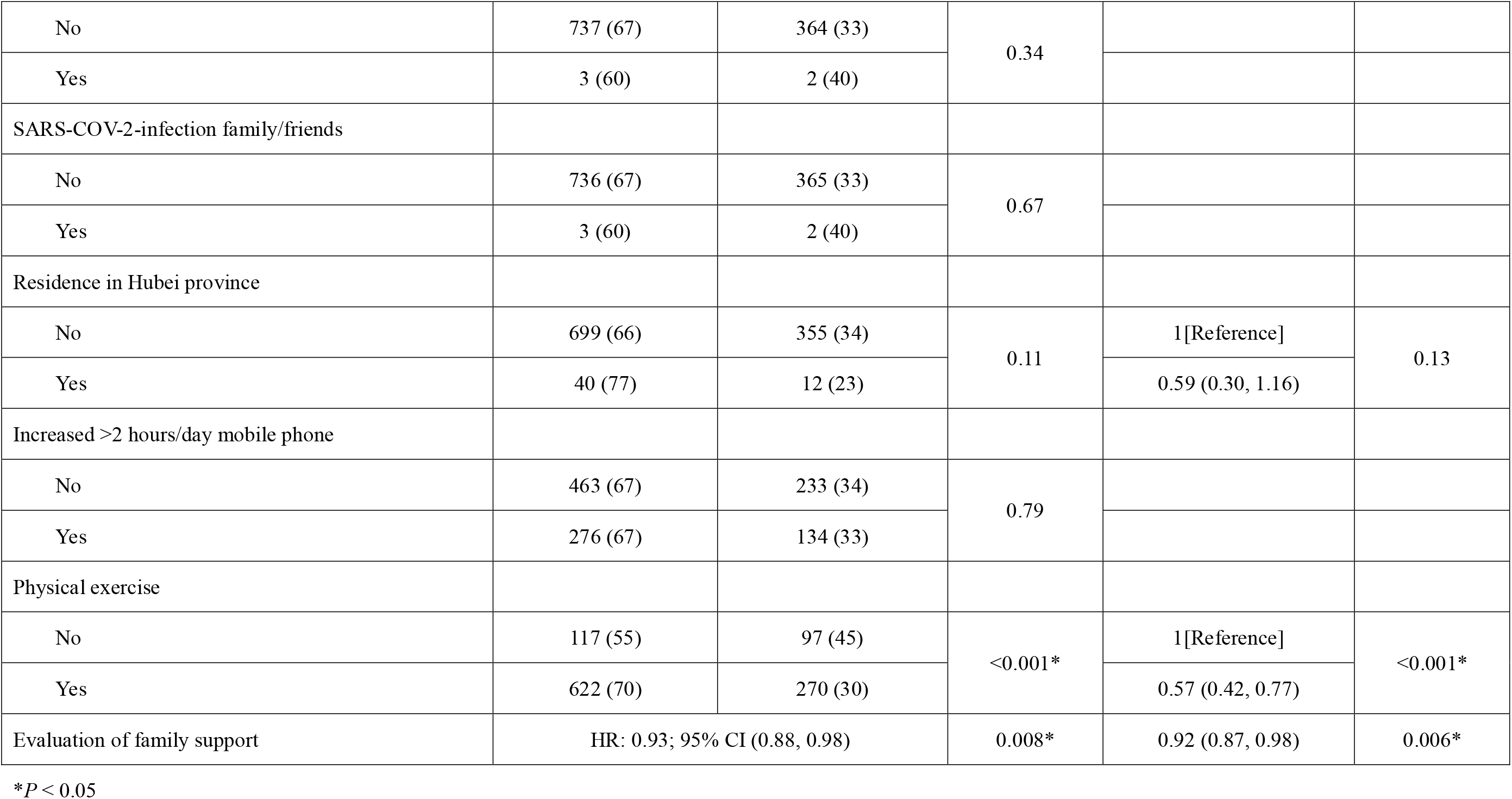
Risk factors of anxiety in lymphoma patient respondents.

### Caregivers

Caregiver respondents were knowledgeable of patients’ diagnoses, stage and therapy, were aware of difficulties patients faced and were enthusiastic to receive information about lymphoma-related medical aspects (Table 1). Patient respondents gave their caregivers a mean support score of 8.84 ± 2.16 (SD).

### Mobile phone use

There was a large increase in mobile phone use during the SARS-CoV-2 pandemic in China. 12% (10, 14%) of patient respondents, 10% (8, 12%) of caregivers and 18% (15, 22%; *P* < 0.001) of normal individuals used their mobile phones (the main source of internet access in China) for ≥ 8 hours *pe*r day *versus* 6% (5, 8%; *P* < 0.001), 5% (4, 6%; *P* < 0.001) and 8% (6, 11%; *P* < 0.001) before the pandemic. 40% (37, 43%), 50% (46, 53%) and 52% (47, 56%) reported an increase in mobile phone use by > 2 hours daily during the pandemic. 86% (83, 88%), 87% (84, 89%), and 90% (87, 92%, *P* = 0.052) of patient, caregiver and normal respondents accessed pandemic news on their mobile phones. 42% (39, 45%), 29% (26, 32%) and 53% (48, 57%; *P* < 0.001) of patient, caregiver and normal respondents reported to use their mobile phone for entertainment including reading novels, movies or TV. 75% (72, 78%) and 84% (81, 86%; *P* < 0.001) of patients and caregiver respondents reported reading news about lymphoma or participated in the CSCO lymphoma education programme.

### Internet-based CSCO programme

1275 (62%) of patients and caregiver respondents reported participating in the CSCO education programme. 236 (35% [31, 38%]) of patient and 238 (40%, [36, 44%], *P* = 0.037) of caregiver respondents participated in ≥ 3 sessions. 53% (49, 57%) of patient and 63% (59, 67%) of caregiver respondents reported increased confidence after participating in the programme (*P* < 0.001). The programme was scored 8.42 ± 1.86 (SD) by patient respondents and 8.47 ± 1.93 by caregiver respondents (*P* = 0.68).

### Quality of life

We used the EORTC QLQ-C30 instrument to compare lymphoma patient respondents’ HRQoL at the time of our study with that of Chinese lymphoma patients in a survey in 2019 after matching for sex, age, education level, lymphoma type and therapy in a propensity score analysis (Additional file 2). Patient respondents had better HRQoL scores compared with controls. Physical, role, emotional, cognitive, social functioning and general HRQoL were significantly better (all *P-*values < 0.05). Nausea and vomiting, dyspnea, insomnia, constipation and financial difficulties were milder (all *P-*values < 0.05) compared with pre-pandemic controls (Table 5).

**Table 5.** HRQoL of lymphoma patients during the pandemic and pre-pandemic.

| HRQoL scale/item | HRQoL of patients, mean (SD) |  | *P*-value |
| --- | --- | --- | --- |
|  | In the pandemic (*n*=1106) | In 2019 pre-pandemic (*n*=1106) |  |
| Global health status/QoL | 70.1 (21.4) | 59.7 (23.1) | <0.001* |
| Functional scale |  |  |  |
| Physical functioning | 81.9 (16.5) | 79.2 (18.1) | 0.025* |
| Role functioning | 81.3 (24.1) | 73.7 (27.3) | <0.001* |
| Emotional functioning | 74.9 (21.0) | 66.0 (23.7) | <0.001* |
| Cognitive functioning | 78.4 (19.6) | 74.4 (21.2) | <0.001* |
| Social functioning | 62.3 (28.6) | 49.3 (30.1) | <0.001* |
| Symptom scale/item |  |  |  |
| Fatigue | 35.9 (22.1) | 41.6 (22.7) | 0.81 |
| Nausea and vomiting | 9.1 (17.2) | 10.7 (18.9) | 0.019* |
| Pain | 18.7 (21.2) | 20.3 (21.5) | 0.12 |
| Dyspnea | 17.1 (21.2) | 22.3 (23.7) | 0.006* |
| Insomnia | 28.6 (29.0) | 31.6 (28.5) | <0.001* |
| Appetite loss | 16.2 (23.3) | 21.2 (24.6) | 0.89 |
| Constipation | 15.1 (5.22) | 16.4 (24.6) | <0.001* |
| Diarrhea | 11.9 (19.3) | 13.8 (20.5) | 0.072 |
| Financial difficulties | 46.3 (34.6) | 62.2 (34.4) | <0.001* |

\* $P < 0.05$

**Table 6.** Co-variates of HRQoL in patient respondents.

| HRQoL scale/item | Co-variables of HRQoL |  |  |  |  |
| --- | --- | --- | --- | --- | --- |
| | Decreased intensity of treatment | Increased >2 hours on mobile phones | Participated in the programs | High evaluation of family support | High evaluation of online project ($n=683$) |
| Global health status/QoL | Worsen<br>(tau b -0.13, $P < 0.001^*$) | No impact<br>(tau b -0.014, $P = 0.60$) | No impact<br>(tau b 0.011, $P = 0.69$) | Improved<br>(tau b 0.12, $P < 0.001^*$) | Improved<br>(tau b 0.15, $P < 0.001^*$) |
| Functional scale |  |  |  |  |  |
| Physical functioning | Worsen<br>(tau b -0.17, $P < 0.001^*$) | No impact<br>(tau b -0.005, $P = 0.85$) | No impact<br>(tau b -0.048, $P = 0.065$) | No impact<br>(tau b 0.026, $P = 0.28$) | Improved<br>(tau b 0.091, $P = 0.003^*$) |
| Role functioning | Worsen<br>(tau b -0.18, $P < 0.001^*$) | No impact<br>(tau b -0.044, $P = 0.12$) | Worsen<br>(tau b -0.067, $P = 0.015^*$) | No impact<br>(tau b -0.020, $P = 0.44$) | Improved<br>(tau b 0.12, $P = 0.001^*$) |
| Emotional functioning | Worsen<br>(tau b -0.12, $P < 0.001^*$) | No impact<br>(tau b -0.034, $P = 0.20$) | No impact<br>(tau b -0.010, $P = 0.78$) | Improved<br>(tau b 0.10, $P < 0.001^*$) | Improved<br>(tau b 0.13, $P < 0.001^*$) |
| Cognitive functioning | Worsen<br>(tau b -0.95, $P < 0.001^*$) | No impact<br>(tau b -0.030, $P = 0.28$) | No impact<br>(tau b -0.011, $P = 0.69$) | Improved<br>(tau b 0.096, $P < 0.001^*$) | Improved<br>(tau b 0.092, $P = 0.004^*$) |
| Social functioning | Worsen<br>(tau b -0.14, $P < 0.001$) | No impact<br>(tau b -0.015, $P = 0.58$) | No impact<br>(tau b -0.039, $P = 0.14$) | No impact<br>(tau b 0.042, $P = 0.096$) | Improved<br>(tau b 0.10, $P = 0.001$*) |
| Symptom scale/item |  |  |  |  |  |
| Fatigue | Worsen<br>(tau b 0.14, $P < 0.001$*) | No impact<br>(tau b -0.008, $P = 0.76$) | No impact<br>(tau b 0.028, $P = 0.28$) | No impact<br>(tau b 0.004, $P = 0.87$) | No impact<br>(tau b -0.035, $P = 0.26$) |
| Nausea and vomiting | Worsen<br>(tau b 0.22, $P < 0.001$*) | No impact<br>(tau b -0.020, $P = 0.48$) | No impact<br>(tau b 0.016, $P = 0.57$) | No impact<br>(tau b -0.013, $P = 0.64$) | Improved<br>(tau b -0.085, $P = 0.011$*) |
| Pain | Worsen<br>(tau b 0.13, $P < 0.001$*) | No impact<br>(tau b 0.027, $P = 0.33$) | No impact<br>(tau b 0.031, $P = 0.26$) | No impact<br>(tau b -0.016, $P = 0.54$) | No impact<br>(tau b -0.041, $P = 0.20$) |
| Dyspnea | Worsen<br>(tau b 0.13, $P < 0.001$*) | No impact<br>(tau b 0.014, $P = 0.64$) | No impact<br>(tau b 0.049, $P = 0.092$) | No impact<br>(tau b -0.050, $P = 0.066$) | Improved<br>(tau b -0.099, $P = 0.004$*) |
| Insomnia | Worsen<br>(tau b 0.069, $P = 0.015$*) | No impact<br>(tau b 0.002, $P = 0.99$) | No impact<br>(tau b 0.009, $P = 0.75$) | Improved<br>(tau b -0.056, $P = 0.033$*) | Improved<br>(tau b -0.12, $P < 0.001$*) |
| Appetite loss | Worsen<br>(tau b 0.22, $P < 0.001$*) | No impact<br>(tau b 0.010, $P = 0.73$) | No impact<br>(tau b 0.052, $P = 0.077$) | No impact<br>(tau b -0.032, $P = 0.24$) | No impact<br>(tau b -0.065, $P = 0.053$) |
| Constipation | Worsen<br>(tau b 0.13, $P < 0.001$*) | No impact<br>(tau b -0.008, $P = 0.80$) | No impact<br>(tau b -0.012, $P = 0.69$) | No impact<br>(tau b -0.033, $P = 0.24$) | No impact<br>(tau b -0.028, $P = 0.41$) |
| Diarrhea | Worsen<br>(tau b 0.10, $P < 0.001^*$) | No impact<br>(tau b 0.009, $P = 0.76$) | No impact<br>(tau b 0.016, $P = 0.58$) | Improved<br>(tau b -0.11, $P < 0.001^*$) | Improved<br>(tau b -0.074, $P = 0.029^*$) |
| Financial difficulties | Worsen<br>(tau b 0.094, $P < 0.001^*$) | No impact<br>(tau b -0.002, $P = 0.93$) | No impact<br>(tau b -0.017, $P = 0.54$) | No impact<br>(tau b -0.022, $P = 0.39$) | No impact<br>(tau b -0.034, $P = 0.28$) |

Next, we analyzed co-variates correlated with HRQoL. Increased daily mobile phone use and participation in the education programme were not correlated with HRQoL. In contrast, reduced therapy intensity was significantly associated with a worse general HRQoL and five worse functions, eight symptoms and financial difficulties. Patient respondents who scored caregiver support high had a better general HRQoL, emotional function, cognitive function and fewer or less severe insomnia and diarrhea. Patient respondents who scored the education programme high had better HRQoL including all five functions and fewer symptoms of nausea and vomiting, dyspnea, insomnia and diarrhea (Table 5).

## Discussion

The SAR-CoV-2 pandemic in China reduced lymphoma patients’ access to medical care including out- and inpatient clinic and hospital visits and direct contact with medical personnel including doctors and nurses. It also decreased potential interactions with other lymphoma patients whom they might encounter in clinic or hospital settings. Because of travel restrictions some patients had to switch their *point-of-contact* for medical care, for example, to a nearby clinic or hospital. Decreased blood testing and pharmacy access led to therapy modifications such as switching to oral drugs. These impacts of the pandemic occurred globally (Chen-See 2020; Triggle).

Because these co-variates are important determinants of anxiety and HRQoL in lymphoma patients we sought to quantify these and to determine the impact, if any, of alternative support resources such as caregivers, patient support organizations, and an online education programme.

There are few studies about anxiety or HRQoL in patients with cancer during the SARS-CoV-2 pandemic. A small study of 77 outpatients with lymphoma in one hospital reported an anxiety incidence of 36% (Romito, et al. 2020). We found the incidence of anxiety in lymphoma patients and caregivers was about 30%, 50% higher than in normal individuals in our survey. In our cross-sectional study more than 70% of respondents had minimal/moderate anxiety. Co-variates associated with low incidence of anxiety included no SARS-CoV-2 infection, not being a lymphoma patient or caregiver, physical exercise, higher education level, a > 2 hour increase in daily mobile phone use and a higher family support score. Amongst patient respondents physical exercise and better caregiver support were associated with less anxiety whereas female sex, receiving therapy and reduced therapy intensity were associated with more anxiety.

Previous cancer patient-caregiver dyads studies reported that caregivers experienced similar or higher anxiety levels compared with patients (Li, et al. 2018; Nipp, et al. 2016; Sklenarova, et al. 2015). We found similarly increased incidence of anxiety in lymphoma patients and caregiver respondents. In China caregivers, typically young family members, are deeply involved in patients’ medical care and related activities based on the concept of filial piety often assuming responsibility for patients’ financial, physical and mental support. A higher proportion of patients with aggressive lymphoma reported by the caregiver compared with patient respondents is consistent with caregivers’ concern for patients. Many studies report that caregivers’ well-being is an important aspect for patients’ mental health (Castillo, et al. 2019). Consequently, we were not surprised to find patients scoring their caregiver support high had a lower incidence of anxiety compared with patients giving low scores.

We were surprised however, to find that HRQoL of lymphoma patients during the pandemic was better than a matched cohort before the pandemic. When we analysed unbalanced co-variates between the two cohorts correlating with HRQoL we found no impact of increased daily mobile phone use. A reduction in therapy intensity, however, was significantly associated with worse HRQoL. Patient respondents who scored caregiver support high had a better general HRQoL, physical and emotional function, cognitive function and fewer or less severe symptoms of insomnia and diarrhea. The lower diarrhea score is presumably related to the perception of severity rather than incidence, frequency, or severity. Social support resources for lymphoma patients besides caregivers included online patient support/discussion groups such as House086 and the CSCO professional education programme. We found subjects who rated the quality of these online tools high had a better HRQoL.

Reducing intensity of cancer therapy during the pandemic is not an evidence-based recommendation (Al-Shamsi, et al. 2020; Di Ciaccio, et al. 2020; Ismael, et al. 2020). However, decreased therapy intensity was reported by 24 percent of patients and caregiver respondents in our survey and was associated with a higher incidence of anxiety and worse HRQoL. Our questionnaire did not allow us to determine why therapy intensity was reduced but limited access to medical care at times when significant resources were used to cope with the challenges of the pandemic is the most likely reason.

There are several limitations of our study. Our survey was online with potential selection biases. For example, our patient respondents were younger than most lymphoma patients, perhaps because of increased internet familiarity and/or access. Our normal cohort had a much younger age than patient and caregiver respondents. Because our survey was cross-sectional it was not possible to compare anxiety and HRQoL in the same respondent before and during the pandemic. In our comparison of HRQoL with a pre-pandemic cohort we were short of data on lymphoma stage so we cannot know if the better HRQoL we observed during the pandemic might result from patients with less advanced lymphoma (Stewart, et al. 2016).

## Conclusion

During the SARS-CoV-2 pandemic lymphoma patients and their caregivers had a significantly higher incidence of anxiety compared with normal individuals. Incidence was increased in persons stopping or educing therapy-intensity. Paradoxically, lymphoma respondents had a better HRQoL compared with pre-pandemic lymphoma controls. Good social support including caregiver support and an online lymphoma education programme were associated with less anxiety and better HRQoL. Reduced therapy intensity was also associated with worse HRQoL. Reduced therapy-intensity in cancer patients during the SARS-CoV-2 pandemic may have negative impact on patient anxiety and HRQoL.

## Data Availability

The datasets used and analyzed during the current study are available from the corresponding author on reasonable request.

## Acknowledgements

Yinan Sun MS, Director of Student Wellness (Yungu School; China) reviewed the questionnaire. Prof. Norbert Schmitz, (University Hospital, Münster, Germany) kindly reviewed the typescript. RPG acknowledges support from the National Institute of Health Research (NIHR) Biomedical Research Centre funding scheme.

## Authors’ contributions

SY designed the study and the questionnaire, analyzed data, interpreted the results and drafted the manuscript. DD conducted the 2019 lymphoma survey and extracted matched subjects from the 2019 lymphoma database. HG conducted the online surveys and collected data in pre- and during the pandemic. RPG helped develop the typescript. JM supervised and organized the CSCO patients education programme and designed the study. XH designed and supervised the study. All authors reviewed and approved the typescript.

## Compliance with ethical standards

### Ethical approval

This study was approved by the Ethical Committee of Peking Univeristy Peoples’ Hospital (Register number 2020PHB173).

### Informed consent

Electronic informed consent was obtained from all respondents who could withdraw at any time during the survey without prejudice.

